# Determinants of tuberculosis incidence in East Asia and Pacific: A panel regression analysis

**DOI:** 10.1101/2020.04.14.20065870

**Authors:** Mark M. Alipio

**Author notes:** Correspondence: Mark M. Alipio. **Declaration of Competing Interests** The author declares no conflict of interest. **Funding** None. No funding to declare.

## Abstract

**Background:** Tuberculosis (TB) remains one of the world’s deadliest communicable disease. To circumvent surges of TB cases, several studies have been carried out analyzing the determinants of TB incidence and recommended policy measures based on the significant indicators. Although the determinants were suggested for strategic planning of TB, the implementation of new measures was either unsuccessful or difficult to realize because of logistical, administrative, and financial constraints. This study aims to unravel potential determinants of TB incidence across 23 countries in East Asia and Pacific. The disentangling of possible association between variables was carried out using panel regression analysis.

**Methods:** This is an ecological multinational-based study utilizing readily accessible public data in the analysis. Carbon dioxide emission, PM2.5 air pollution exposure, unemployment (percent of total labor force), percent of people using at least basic sanitation services, percent of people practicing open defecation, health expenditure (percent of GDP), and out-of-pocket health expenditure are included as the determinants of TB incidence. The single outcome variable of this study was TB incidence which is the estimated number of new and relapse tuberculosis cases arising in a given year, expressed as the rate per 100,000 population. A total of 23 countries in the East and Pacific region were included as sampling unit with a time-series length of five years (2010 – 2014), producing 115 samples. Given the nature of data, a panel regression was used to estimate the relationship between the potential determinants and TB incidence.

**Findings:** A significant regression coefficient was found (F(7,107) = 37.9, p < 0.05) with R^2^ = 0.7126. The R^2^ value suggested that 71.26% of the variance in TB incidence was accounted for by the variables in this study. For every one unit increase in microgram per cubic meter of PM2.5 pollution, in the unemployment percentage of total labor force, and in the percentage of out-of-pocket health expenditure, the rate of TB cases per 100,000 population was predicted to be 4.617, 13.504, and 3.467 higher, respectively, holding other variables constant. On the other hand, for every one unit increase in the kiloton of CO_2_ emission and in the percent of people using at least basic sanitation services, the rate of TB cases per 100,000 population was predicted to be 0.00003828 and 4.457 lower, respectively. Percent of people practicing open defecation and health expenditure (percent of GDP) did not significantly influence TB incidence.

**Interpretation:** The countries in the East Asia and Pacific with low PM2.5 air pollution exposure, low unemployment, low out-of-pocket health expenditure, high carbon dioxide emission and high percent of people using at least basic sanitation service, had low incidence of TB for the five-year period. The study suggests how an increase in unemployment consequently increases TB incidence across the countries. Proper implementation of programs that could promote proper hygiene is essential to increase adherence of people to basic sanitation practices. Based on the study, this is an important factor in mitigating higher incidence of TB. Therefore, strategies may be formulated to either maintain or improve this determinant in order to significantly reduce TB cases. Finally, concerted efforts may be developed to decrease emission of hazardous finer particles from residential, industrial, and agricultural burning, in order to control tuberculosis.

## INTRODUCTION

Tuberculosis (TB) is an infectious disease caused by a bacterium called *Mycobacterium tuberculosis*. It is transmitted from a TB patient to another person through coughing, sneezing and spitting. Thus, close contacts, especially household members, could be infected with TB. Lungs are commonly affected but it could also affect other organs such as the kidney, bones, liver and others. TB is curable and preventable. However, incomplete and irregular treatment may lead to drug-resistant TB or death (Franks, Hirsch-Moverman & Colson, 2015).

TB affects mankind for several centuries and still a current major concern of in the field of public health worldwide, despite the strategies employed for effective tuberculosis control. Global Tuberculosis report 2014 states that tuberculosis remains one of the world’s deadliest communicable disease. In 2013, an estimated 9 million people developed TB and 1.5 million died from the disease. Tuberculosis ranks as the second leading cause of death among infectious diseases worldwide (Global Tuberculosis Report, 2014). With these, scenario, tuberculosis remains a major global health problem.

Since 1993, World Health Organization (WHO) has recommended strategy through which national governments can meet their responsibility to treat patients and to prevent the spread of tuberculosis. Four major elements of the strategy, which came to be known as DOTS (Direct Observe Treatment Short Course), were political commitment by governments, improved laboratory services, a continuous supply of good quality drugs and, a reporting system to document the progress (and failure) of treatment for individual patients to complete treatment. To put it simply, direct observation of treatment is an integral and essential component of DOTS.

WHO has reported that more than 30 million patients with tuberculosis have been treated with its five-element DOTS strategy, resulting in cure rates of >80% and default rates of <10% (WHO Bulletin, 2015). WHO’s Global Plan to stop TB highlights the need to expand DOTS through standardized treatment, under proper case management conditions, including directly observed treatment to reduce the risks of acquiring drug resistance, and support of patients to increase adherence to treatment and chance of cure.

In 2010, according to DOH as stated by Ong (2012) tuberculosis is the sixth leading cause of mortality with a rate of 73 Filipinos dying every day of TB and accounts for 5.1 % total of deaths in the Philippines. Out of 196 countries, the Philippines has the distinction of being included in the Top 22 high-burden tuberculosis countries in the world, which ranks the Philippines number nine worldwide (WHO, 2012). Together, these 22 countries (Including the Philippines) contribute to 80 percent of global TB burden.

The WHO South-East Asia (SEA) Region is home to 26% of the world’s population with 44% burden of TB incidence. In 2017, an estimated 4.4 million people fell ill with TB and estimated 638,000 died because of the disease which is more than half of global TB deaths. Treatment success for new and relapse TB cases was 75% (for those initiated on treatment in 2016), amongst the lowest in the Regions of the world. It is also estimated that 192 000 Rifampicin-resistant (RR) and multi-drug-resistant TB (MDR-TB) cases accounting for more than 34% of global burden appeared in the Region in 2017, of which less than 52 000 were notified in the same year. Six out of the 30 high TB (and MDR-TB) burden countries are in the SEA Region: Bangladesh, Democratic People’s Republic of Korea, India, Indonesia, Myanmar and Thailand.

The Philippine government addresses the problem of TB in the country through the National Tuberculosis Program (NTP). The NTP is one of the public health programs being managed and coordinated by the infectious Disease for health programs being managed and coordinated Infectious Disease for the Prevention and Control Division of the Diseased Prevention and Control Bureau (DPCB) of the DOH. The NTP has the mandate to develop TB control policies, standards and guidelines, formulate the national strategic plan, manage program logistics, provide leadership and technical assistance to the lower health offices/units, manage data, and monitor and evaluate the program. The program’s TB diagnostic and treatment protocols and strategies are in accordance with the global strategy of STOP TB partnership and the policies of the World Health Organization (WHO) and the International Standards for TB Care (ISTC).

The overarching strategy of the NTP is the DOTS or Direct Observed Treatment Short Course that was started in the country in 1996. This was expanded under the WHO-endorsed STOP TB strategy that the country adopted from 2006-2010. In, 2010, DOH issued the 2010-2016 Philippine Plan of Action to Control TB (PhilPACT) as the roadmap for controlling TB.

However, Yadav et al. (2010) states that not all TB patients seek medical care through the National TB program. A large segment of the population seeks healthcare from the private sector that includes from the private sector that includes private medical physicians, paramedics, physicians from private hospitals and nursing homes, non-government organizations (NGO’S) and the corporate sector health-care institutions. To reach out these TB patients, the NTP adopted the Public-Private Mixed DOTS (PPMD) strategy in 2003 to increase case detection and harmonize TB management among all healthcare providers in the country. Around 6% of the total TB cases nationwide were contribute by this initiative in 2008 (NTCCP-MPO 2014). Presently, the Philippines’ DOTS treatment success is at around 88%, which is higher that the WHO target at 85% (Ong, 2012).

Despite the implemented strategies, TB remains a burden in several countries across the world. Several studies have been carried out analyzing the determinants of TB incidence and recommended policy measures based on the significant indicators. In African and American countries, the incidence of TB decreases quickly with higher health expenditure, higher income, lower immigration, lower child mortality, and lower HIV infection rate (Dye, Lönnroth, Jaramillo, Williams, & Raviglione, 2009). In Europe, TB incidence variation is explained by a country’s level of egalitarianism and wealth (Ploubidis et al., 2012). Social protection and urban planning were found to strengthen tuberculosis control (Hargreaves et al., 2011). AIDS incidence rate, unemployment rate, Gini coefficient, proportion of inmates, mean per capita household income, and primary care coverage were significant determinants of TB incidence (Pelissari, & Diaz-Quijano, 2017). Although the determinants were suggested for strategic planning of TB, the implementation of new measures was either unsuccessful or difficult to realize because of logistical, administrative, and financial constraints (Dye et al., 2009; Hargreaves et al., 2011).

### POTENTIAL DETERMINANTS OF TB INCIDENCE

#### Carbon dioxide (CO_2_) Emission

CO_2_ is a colorless and odorless gas formed by the combustion of carbon and in the respiration of living organisms. It is considered a greenhouse gas. Emissions of CO_2_ are those curtailing from fossil fuels burning and the manufacture of cement. They include CO_2_ produced during consumption of solid, liquid, and gas fuels and gas flaring. Existing evidence showed CO_2_ influences resistance of TB suggesting how higher amounts of CO_2_ could affect treatment outcome of TB patients (Corper, Gauss, & Rensch, 1921; Schaefer, Cohn, & Middlebrook, 1955). In this study, CO_2_ emission is expressed in kilotons.

#### PM2.5 Air Pollution Exposure

PM2.5 refers to particulate matter (PM) in the atmosphere with a less than 2.5 microns diameter, which is about 3% the diameter of a human hair. The particles come from several sources including residential wood burning, power plants, motor vehicles, volcanic eruptions, dust storms, forest fires, agricultural burning, and airplanes. Population-weighted exposure to ambient PM2.5 pollution is the mean exposure level of a country’s population to concentrations of suspended particles which could penetrate deep to the respiratory system and cause severe health damage. The exposure is computed by weighting mean yearly concentrations of PM2.5 by population in urban and rural communities. Studies have shown that exposure to PM2.5 could trigger cardiovascular disease-related mortality and respiratory problems (Dockery & Stone, 2007; Xing, Xu, Shi, & Lian, 2016). Scientists in the study estimated that for every 10 μg/m^3^ increase in fine PM2.5 pollution, there is an associated 4%, 6% and 8% increased risk of all-cause, cardiopulmonary and lung cancer mortality, respectively (Pope et al., 2002). In this study, the mean annual exposure is expressed in micrograms per cubic meter.

#### Unemployment (Percent of Total Labor Force)

The World Bank defined unemployment as the “share of the labor force that is without work but available for and seeking employment.” Previous studies have demonstrated unemployment as one of the strongest determinants of infection incidence in several countries across the world. Among the infections intertwined with unemployment were HIV, AIDS, and several respiratory problems (Grob et al., 2016; Hunter, 2007). The studies showed how an increase in unemployment consequently increases infection rate.

#### Sanitation and Defecation Practices

The percentage of people using at least basic sanitation services, that is, improved sanitation facilities that are not shared with other households. This indicator encompasses both people using basic sanitation services as well as those using safely managed sanitation services. Improved sanitation facilities include flush/pour flush to piped sewer systems, septic tanks or pit latrines; ventilated improved pit latrines, compositing toilets or pit latrines with slabs. People practicing open defecation refers to the percentage of the population defecating in the open, such as in fields, forest, bushes, open bodies of water, on beaches, in other open spaces or disposed of with solid waste.

#### Health Expenditures

Current health expenditure (percent of GDP) is the level of current health expenditure expressed as a percentage of GDP. Estimates of current health expenditures include healthcare goods and services consumed during each year. This indicator does not include capital health expenditures such as buildings, machinery, IT and stocks of vaccines for emergency or outbreaks. Share of out-of-pocket payments of total current health expenditures. Out-of-pocket expenditure (percent of current health expenditure) is the out-of-pocket payment that are spent on health directly out-of-pocket by households.

### RELATED LITERATURE

#### Government Healthcare Facility and Private Healthcare Facility

These are two different entities that shared one common goal – eradicate and help the people against TB (Manalaysay, 2014). They have its common goal but they do differ in terms with the source of its allocations and funding. The Government healthcare facilities provides treatment to the patient that is free of charge. On the other hand, the private healthcare facility is an institution that treat illness with a corresponding payment due to its business venture.

In public administration there is an intimate relationship between service rendered and the cost of the service charged from the public. Only such amount of money is raised by taxation, which is necessary for the rendering of service. In private administration income of funds exceeds expenditure because there is usually an attempt to extract as such money as possible from the public (Paul, 2011).

The research carried out by Peters & Savoie (2011) explains that “the tasks of governing are almost inherently more difficult than the tasks of managing in the private sector, given the multiple goals, the constraints on action, and the demands for accountability that characterize the public sector”.

#### Role of Public Administration

According to Maas (1959), editor of Area and Power, local government is presented as a manner of dividing power by area or authority. He pointed out that the creation of local government units is “advantageous for the promotion of rural development”. Local governments are a means of providing self-identity, especially in ethnically homogenous communities. This would be of greater help to support the community in realizing the needs of their constituents. In the Philippines, the local government establishes rural healthcare facilities to cater the needs in terms of health among the citizens. This is to address the universal access to health among Filipinos.

According to Chand and Chakrabarty (2012), since the birth of public administration, its study has been growing in different directions and involves complex concerns and functions. Thus, in an attempt to define the different aspects of public administration, these are described in relatively distinct approaches that grow out of the different perspectives that shape its structures and functions. Each approach gives a particular point of view of administrative activity. These different approaches are best regarded as ways in which to approach the health issues in a specific country.

### FRAMEWORK

#### Theoretical Framework

The study is anchored on the theory of Health Belief Model of Becker (1978). The health belief model is a psychological health behavior change model developed to explain and predict health-related behavior, particularly in regard to the uptake of health services. This model suggests that people’s belief about health problems, perceived benefit of action and barriers to action, and self-efficacy explain engagement in health promoting behavior. A stimulus, or cue to action, must also be present in order to trigger the health-promoting behavior.

The model is relevant in the study since the model believes that given appropriate interventions, individuals will be reminded and encouraged to engage in health promoting behaviors. These behaviors can be aimed at the individual level or the social level.

Moreover, this study was anchored on Lydia Hall’s “Care, Cure, Core Theory. According to the theory, nurses are focused on performing the noble task of nurturing the patients. This is represented by the care circle that talks about the role of nurses, and is focused is nurturing the patient meet nay need he or she unable to meet alone.

Meanwhile, the core circle represents the patient receiving the nursing care. The core has goals set by him or himself rather than by any other person, and behaves according to his or her feelings and values. This involves the therapeutic use of self, and is shared with other members of the health team.

On the other hand, the cure circle explains the aspects of nursing which involves the administration of medications and treatments. Hall explains in the model that the cure circle is shared by the nurse with other health professionals such as physicians or physical therapists.

Applying Lydia Hall’s Theory to this study, the interventions or actions enumerated above geared towards treating the patient for whatever illness or disease he or she is suffering from. Thus, the results of the study would assist the TB DOTS nurse to be an active advocate of the patient thereby helping the patient to be cured or complete the TB DOTS program.

Smith (2007) argued to equate governance with government means to focus on technical problems of administrative and legal capacity and the improvement of public sector management, the legal framework for development, accountability through better auditing, decentralization, the policing of corruption, civil service reform, and improved information on policy issues for both decision-makers and the public. There are critics to this approach for its’ ‘managerialist fixes’, ‘detached from the turbulent world of social forces, politics and the structure and purpose of the state’. In this context it is important to recognize the support for civil society in order to encourage political accountability, legitimacy, transparency and participation.

#### Conceptual Framework

As can be seen in the conceptual paradigm (**Figure 1**), carbon dioxide emission, PM2.5 air pollution exposure, unemployment (percent of total labor force), percent of people using at least basic sanitation services, percent of people practicing open defecation, health expenditure (percent of GDP), and out-of-pocket health expenditure are included as the determinants of TB incidence.

**Figure 1.**
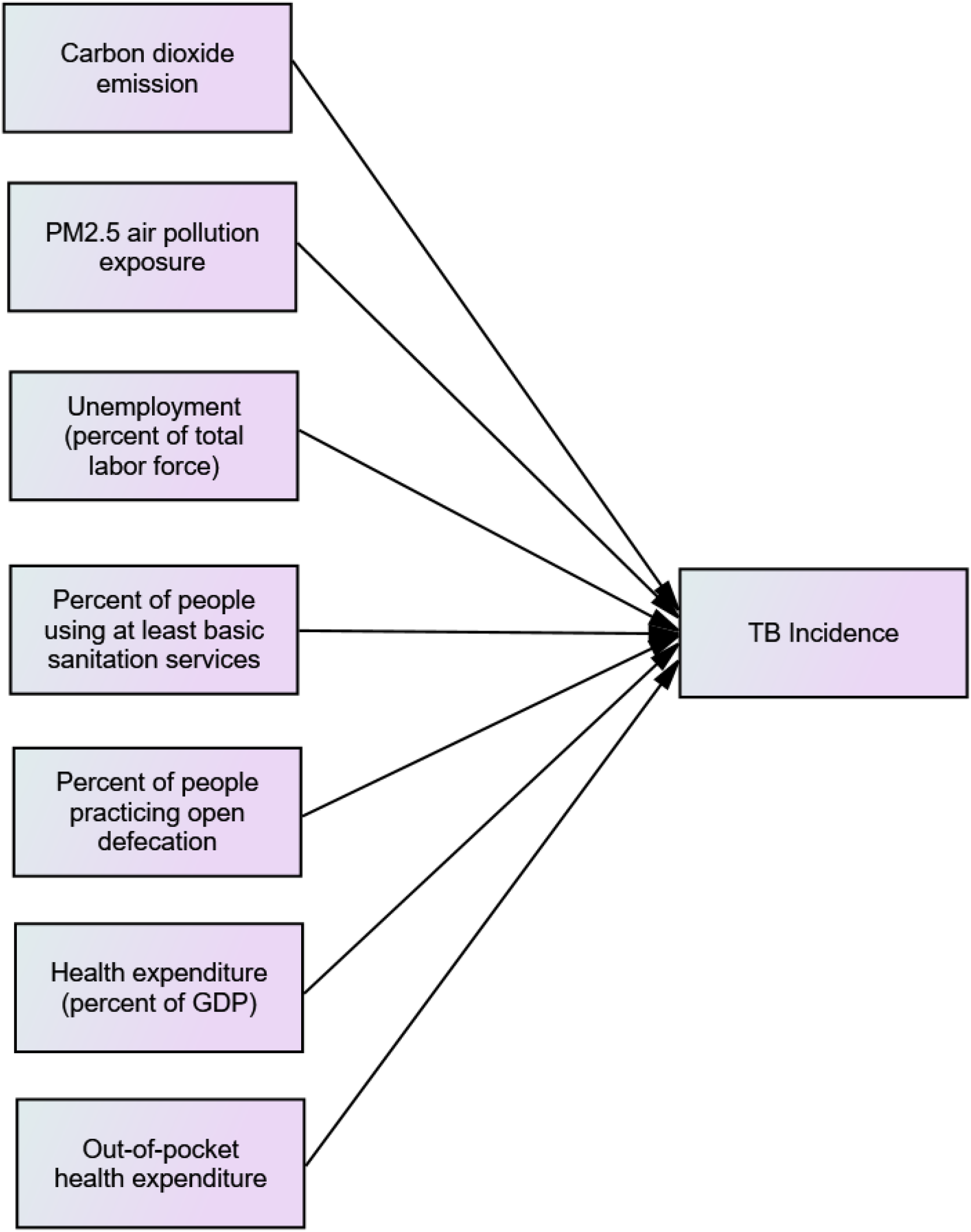
Conceptual paradigm.

It is hypothesized that percent of people using at least basic sanitation services, health expenditure (percent of GDP), and out-of-pocket health expenditure would positively influence TB incidence. Carbon dioxide emission, PM2.5 air pollution exposure, unemployment (percent of total labor force), and percent of people practicing open defecation would negatively influence TB incidence.

## METHODOLOGY

This is an ecological multinational-based study that was conducted using data from the World Bank Open Data (https://data.worldbank.org/) on tuberculosis incidence and its potential determinants. The data pertaining to the variables under study were extracted retrospectively. Trends for the period 2010 – 2014 were considered. Only a five-year analysis was conducted due to missing data in several determinants employed in the study. Carbon dioxide emission, PM2.5 air pollution exposure, unemployment (percent of total labor force), percent of people using at least basic sanitation services, percent of people practicing open defecation, health expenditure (percent of GDP), and out-of-pocket health expenditure are included as the determinants of TB incidence. The single outcome variable of this study was TB incidence which is the estimated number of new and relapse tuberculosis cases arising in a given year, expressed as the rate per 100,000 population. All forms of TB are included, including cases in people living with HIV. Estimates for all years are recalculated as new information becomes available and techniques are refined, so they may differ from those published previously.

A total of 23 countries in the East and Pacific region were included as sampling unit with a time-series length of five years, producing 115 samples. Given the nature of data, a panel regression was used to estimate the relationship between the potential determinants and TB incidence. This analysis is commonly used in econometrics, where the behavior of panel units is followed across time. Before regressing, variables were transformed into log form to have a more straightforward interpretation and comparison of the size of the estimated coefficients and to secure the assumption of regression on heteroscedasticity. Also, assumptions on the normality of residuals, no collinearity relation between independent variables and no serial autocorrelation of errors were diagnostically checked in this paper. All analyses were carried using R statistical package.

The following equation expresses TB incidence model for this study:

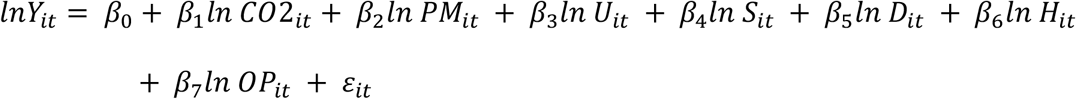

where:

lnY_it_ = log TB Incidence for country “i” at time “t”

ln CO2_it_ = log Carbon dioxide emission for country “i” at time “t”

ln PM_it_ = log PM2.5 air pollution exposure for country “i” at time “t”

ln U_it_ = log Unemployment for country “i” at time “t”

ln S_it_ = log % people using basic sanitation services for country “i” at time “t”

ln D_it_ = log % people practicing open defecation for country “i” at time “t”

ln H_it_ = log health expenditure for country “i” at time “t”

ln OP_it_ = log outofpocket health expenditurefor country “i” at time “t”

ε_it_ = random error

β_0_, β_1_, β_2_, β_3_, β_4_, β_5_, β_6_, β_7_ = study parameters

### RESULTS AND DISCUSSION

### Diagnostic Checking

Several assumptions for running the panel regression analysis were checked for suitability of the model. It is essential to evaluate the aptness of the generated model for the said data and testing it to see whether it satisfies the required assumptions for its appropriateness. The data analysis utilized the different formal tests and generated the results from R software.

In the figure below, the relationship between the predictors and TB incidence is considerably linear.

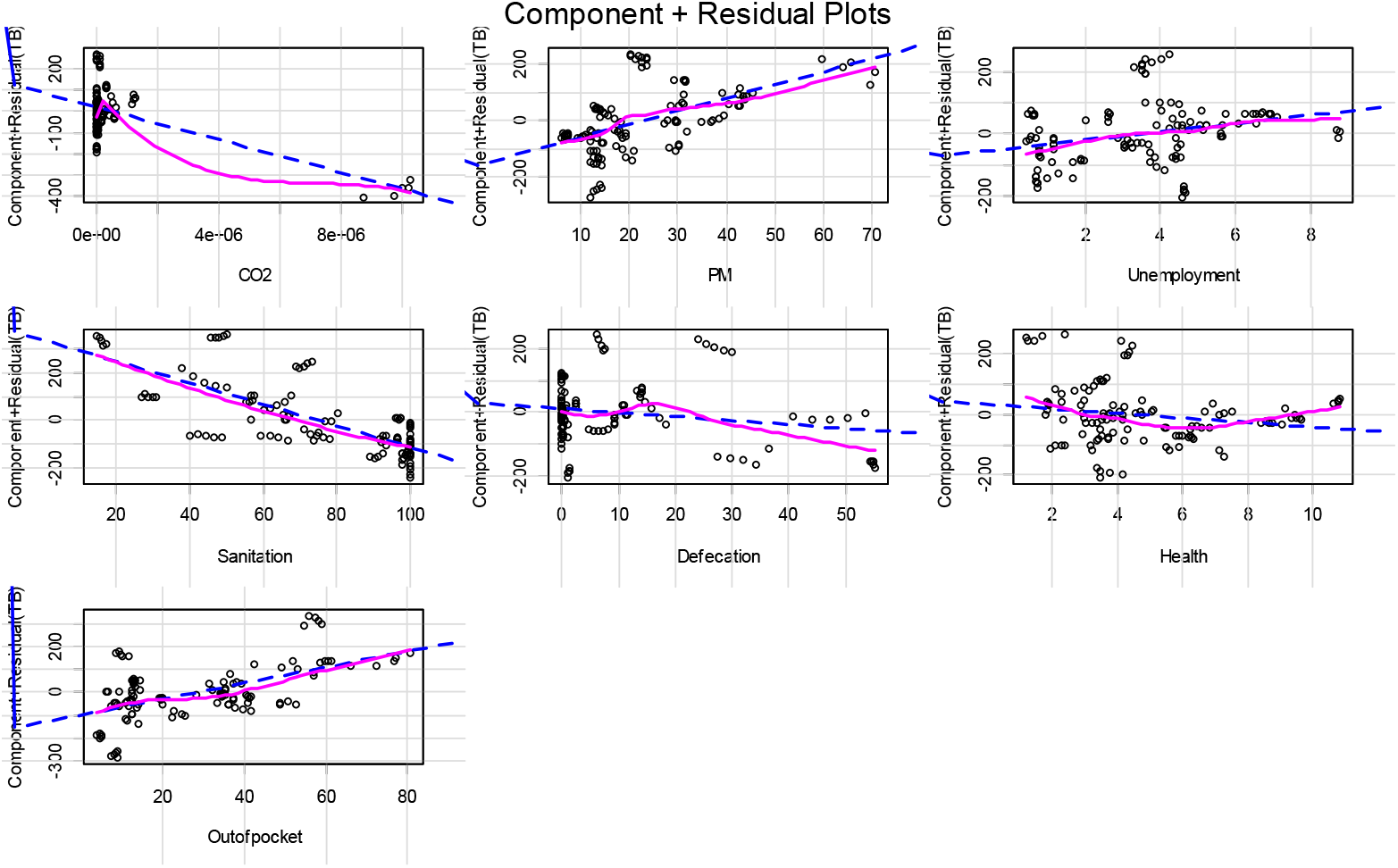

The following were the Variance Inflation (VIF) values of the predictors: Carbon dioxide emission = 2.51, PM2.5 air pollution exposure = 3.69, unemployment (percent of total labor force) = 1.72, percent of people using at least basic sanitation services = 2.58, percent of people practicing open defecation = 2.66, health expenditure (percent of GDP) = 1.27, and out-of-pocket health expenditure = 1.80. All values lie within 1-10 indicating that there is no multicollinearity. In other words, all of the predictors are not too highly correlated with each other.

Durbin-Watson (D-W) test calculated a D-W statistic of 1.34. As a general rule of thumb, the residuals are uncorrelated when the Durbin-Watson statistic is approximately 2. In this case, the residuals for the model are uncorrelated indicating that the values of the residuals are independent.

To check for homoscedasticity, the Breush-Pagan and Goldfeld-Quandt tests were employed. The null hypothesis for Breush-Pagan and Goldfeld-Quandt tests assumes homoscedasticity and a p-value below a certain level (0.05) indicates that the null hypothesis should be rejected in favor of heteroscedasticity. Both tests were not significant (BP = 22.602, df = 7, p-value = 0.211999; GQ = 1.4459, df1 = 50, df2 = 49, p-value = 0.09938) indicating that the variance of the residuals is constant.

To identify if there are influential cases biasing the model, the studentized residuals with Bonferroni was used. The test revealed that there are no potential outliers of the model (r-student = 2.57, unadjusted p-value = 0.12, Bonferroni p = NA).

### TB Incidence Model

A panel linear regression was calculated to predict TB incidence (Y) based on carbon dioxide emission (CO_2_), PM2.5 air pollution exposure (PM), unemployment (percent of total labor force) (U), percent of people using at least basic sanitation services (S), percent of people practicing open defecation (D), health expenditure (percent of GDP) (H), and out-of-pocket health expenditure (OP) (**Table 1**). A significant regression coefficient was found (F(7,107) = 37.9, p < 0.05) with R^2^ = 0.7126. The R^2^ value suggested that 71.26% of the variance in TB incidence was accounted for by the variables in this study. The generated model is shown in the next section.

**Table 1.** Parameter estimates of the model

| Parameters | Unstandardized Coefficients |  | Standardized Coefficients | t | Sig. |
| --- | --- | --- | --- | --- | --- |
|  | B | Std. Error | Beta |  |  |
| (Constant) | 340.444 | 64.630 |  | 5.268 | .000 |
| CO<sub>2</sub> emission | -3.828E-5 | .000 | -.420 | -5.119 | .000 |
| PM<sub>2.5</sub> air pollution | 4.617 | 1.301 | .353 | 3.549 | .001 |
| Unemployment | 13.504 | 5.916 | .155 | 2.283 | .024 |
| Sanitation | -4.457 | .596 | -.623 | -7.480 | .000 |
| Defecation | -1.261 | .993 | -.107 | -1.270 | .207 |
| Health Expenditure | -7.612 | 4.447 | -.100 | -1.712 | .090 |
| Out-of-pocket | 3.467 | .646 | .373 | 5.367 | .000 |

### Model

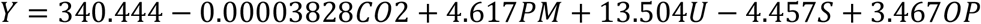

where:

Y = TB incidence

CO2 = Carbon dioxide emission

PM = PM2.5 air pollution exposure

U = Unemployment (percent of total labor force)

S = Percent of people using at least basic sanitation services

OP = Out − of − pocket health expenditure

**Note:** Percent of people practicing open defecation (D) and health expenditure (percent of GDP) (H) are not included in the model since they are not significant.

PM2.5 air pollution exposure (PM), unemployment (percent of total labor force) (U), and out-of-pocket health expenditure (OP) were positively associated with TB incidence (Y); however, carbon dioxide emission (CO_2_) and percent of people using at least basic sanitation services (S) were negatively associated with TB incidence. The findings suggest that increasing PM2.5 air pollution exposure, unemployment, and out-of-pocket health expenditure are associated with increasing TB incidence; while increasing carbon dioxide emission and percent of people using at least basic sanitation services are associated with decreasing TB incidence. Conversely, decreasing PM2.5 air pollution exposure, unemployment, and out-of-pocket health expenditure are associated with decreasing TB incidence; while decreasing carbon dioxide emission and percent of people using at least basic sanitation services are associated with increasing TB incidence. Meanwhile, percent of people practicing open defecation and health expenditure (percent of GDP) do not significantly influence TB incidence.

The parameter estimates of the model are worthwhile to discuss. For every one unit increase in microgram per cubic meter of PM2.5 pollution, in the unemployment percentage of total labor force, and in the percentage of out-of-pocket health expenditure, the rate of TB cases per 100,000 population is predicted to be 4.617, 13.504, and 3.467 higher, respectively, holding other variables constant. On the other hand, for every one unit increase in the kiloton of CO_2_ emission and in the percent of people using at least basic sanitation services, the rate of TB cases per 100,000 population is predicted to be 0.00003828 and 4.457 lower, respectively. When kiloton is converted to kilogram, this implies that for every one unit increase in the kilogram of CO_2_ emission, the rate of TB cases per 100,000 population is predicted to be 38.28 lower.

The findings revealed that the countries in the East Asia and Pacific with high PM2.5 air pollution exposure, high unemployment, and high out-of-pocket health expenditure, had high incidence of TB for the five-year period. The countries with low carbon dioxide emission and low percent of people using at least basic sanitation service, had high incidence of TB.

The findings also revealed that the countries in the East Asia and Pacific with low PM2.5 air pollution exposure, low unemployment, and low out-of-pocket health expenditure, had low incidence of TB for the five-year period. The countries with high carbon dioxide emission and high percent of people using at least basic sanitation service, had low incidence of TB.

This result suggested that higher mean exposure level of a country’s population to concentrations of suspended particles which could penetrate deep to the respiratory system and cause severe health damage is associated with higher incidence of TB. Empirical evidence showed that exposure to PM2.5 could trigger cardiovascular disease-related mortality and respiratory problems (Dockery & Stone, 2007; Xing, Xu, Shi, & Lian, 2016). Scientists in one study estimated that for every 10 μg/m^3^ increase in fine PM2.5 pollution, there is an associated 4%, 6% and 8% increased risk of all-cause, cardiopulmonary and lung cancer mortality, respectively (Pope et al., 2002).

One notable finding of this research indicated that the higher the share of the labor force that is without work but available for and seeking employment in a given country, the higher is the incidence of TB. Previous studies have demonstrated unemployment as one of the strongest determinants of infection incidence in several countries across the world. Among the infections intertwined with unemployment were HIV, AIDS, and several respiratory problems (Grob et al., 2016; Hunter, 2007). The studies showed how an increase in unemployment consequently increases infection rate.

It can be gleaned that lower out-of-pocket health expenditure is associated with low incidence of TB. The out-of-pocket health expenditures are the expenses that the patient or the family pays directly to the healthcare provider, without a third-party (insurer, or State). The lower are the direct payments made by individuals to healthcare providers at the time of service use, the lower is the incidence of TB.

It can also be noted that the higher is the percent of people using at least basic sanitation services, the lower is the incidence of TB. Improved sanitation facilities such as flush/pour flush to piped sewer systems, septic tanks or pit latrines; ventilated improved pit latrines, compositing toilets or pit latrines with slabs, are associated with low TB incidence among the countries. This highly suggests how proper sanitation practices are essential in mitigating higher incidence of TB.

## CONCLUSION

In conclusion, the countries in the East Asia and Pacific with low PM2.5 air pollution exposure, low unemployment, low out-of-pocket health expenditure, high carbon dioxide emission and high percent of people using at least basic sanitation service, had low incidence of TB for the five-year period. The study suggests how an increase in unemployment consequently increases TB incidence across the countries. Proper implementation of programs that could promote proper hygiene is essential to increase adherence of people to basic sanitation practices. Based on the study, this is an important factor in mitigating higher incidence of TB. Therefore, strategies may be formulated to either maintain or improve this determinant in order to significantly reduce TB cases. Finally, concerted efforts may be developed to decrease emission of hazardous finer particles from residential, industrial, and agricultural burning, in order to control tuberculosis.

## Data Availability

The author confirms that the data supporting the findings of this study are available within the article and its supplementary materials.

https://data.worldbank.org/

## Notes

### Competing Interest Statement

The authors have declared no competing interest.

## REFERENCES

Chand, P. and Chakrabarty, B.(2012), Public Administration in the Globalizing World: Theories and Practices.

Corper, H. J., Gauss, H., & Rensch, O. B. (1921). Studies on the Influence of Carbon Dioxide on Resistance to Tuberculosis: The Effect of Carbon Dioxide on the Tubercle Bacillus. American Review of Tuberculosis, 5(7), 562–587.

Dockery, D. W., & Stone, P. H. (2007). Cardiovascular risks from fine particulate air pollution. N Engl J Med, 356(5), 511–513.

Dye, C., Lönnroth, K., Jaramillo, E., Williams, B. G., & Raviglione, M. (2009). Trends in tuberculosis incidence and their determinants in 134 countries. Bulletin of the World Health Organization, 87, 683–691.

Alipio, M. (2020). Do Latitude and Ozone Concentration Predict COVID-2019 Cases in 34 Countries?. Available at SSRN 3572114.

Alipio, M. (2020). Vitamin D Supplementation Could Possibly Improve Clinical Outcomes of Patients Infected with Coronavirus-2019 (COVID-2019). Available at SSRN 3571484.

Alipio, M. (2020). Epidemiology and Clinical Characteristics of 50 Death Cases with COVID-2019 in the Philippines: A Retrospective Review. Available at SSRN 3570612.

Getahum B., Amen, G., Medhin, G., Biadgilin, S. (2010). Treatment outcome of tuberculosis patients under directly observed treatment in Addis, Ababa, Ethopia. Retrieved from

Global Tuberculosis Report 2014 (2014). Retrieved last November 12, 2018 from http://www.who.int/tb/publications/global_report/gtbr14_main_

Grob, M., Herr, A., Hower, M., Kuhlmann, A., Mahlich, J., & Stoll, M. (2016). Unemployment, health, and education of HIV-infected males in Germany. International journal of public health, 61(5), 593–602.

Hargreaves, J. R., Boccia, D., Evans, C. A., Adato, M., Petticrew, M., & Porter, J. D. (2011). The social determinants of tuberculosis: from evidence to action. American journal of public health, 101(4), 654–662.

Alipio, M. M. (2020). Academic Adjustment and Performance among Filipino Freshmen College Students in the Health Sciences: Does Senior High School Strand Matter.

Hunter, M. (2007). The changing political economy of sex in South Africa: The significance of unemployment and inequalities to the scale of the AIDS pandemic. Social science & medicine, 64(3), 689–700.

Life experiences of patients who have completed tuberculosis treatment: a qualitative investigation in Southeast Brazil. Httpp://www.biomedcentral.com/14712458/13/595

Lima Dias, A. Falcao de Oliveria, D., Turato, E., Moralez de Figueirido R. (2013). Maas, Arthur (1959), ed. Area and Power: A Theory of Local Government Glencoe, III.: Free Press.

Alipio, M. M. (2020). Chest Radiographic Findings of Patients Infected with 2019-nCOV. Chest.

Manalaysay, C. (2014). The Scope of Government health services and private health services.

Nissen, T., Rose, M., Kimaru, G., Bygbjerg, I., Mfinanga, S., Ravn, P. (2012). Challenges of Loss to Follow-up in Tuberculosis Research.

Ong, Willie (2012). Tuberculosis in the Philippines : 10 things you should know. http://www.philstar.com.ph

Paul, S. (2011). Public Administration: Theory and Practice. University of Calicut. School of Distance Education. Core course Module

Alipio, M., & Pregoner, J. D. (2020). Epidemiological Characteristics of An Outbreak of Coronavirus Disease 2019 in the Philippines. Available at SSRN 3568934.

Pelissari, D. M., & Diaz-Quijano, F. A. (2017). Household crowding as a potential mediator of socioeconomic determinants of tuberculosis incidence in Brazil. PLoS One, 12(4).

Peters, B. Guy, Savoie, D. (2011). Governance in the Twenty-first Century.

Alipio, M. M. (2020). Predicting Academic Performance of College Freshmen in the Philippines using Psychological Variables and Expectancy-Value Beliefs to Outcomes-Based Education: A Path Analysis.

Ploubidis, G. B., Palmer, M. J., Blackmore, C., Lim, T. A., Manissero, D., Sandgren, A., & Semenza, J. C. (2012). Social determinants of tuberculosis in Europe: a prospective ecological study. European Respiratory Journal, 40(4), 925–930.

Pope Iii, C. A., Burnett, R. T., Thun, M. J., Calle, E. E., Krewski, D., Ito, K., & Thurston, G. D. (2002). Lung cancer, cardiopulmonary mortality, and long-term exposure to fine particulate air pollution. Jama, 287(9), 1132–1141.

Revitalizing the Public Service, Canadian Centre for Management Development/ McGill-Queen’s University Press.

Schaefer, W. B., Cohn, M. L., & Middlebrook, G. (1955). The roles of biotin and carbon dioxide in the cultivation of Mycobacterium tuberculosis. Journal of bacteriology, 69(6), 706.

Smith, B.C. (2007), Good Governance and Development, Palgrave Macmillan. Text.pdf

Alipio, M. (2020). A Structural Model of Organizational Commitment among Higher Education Economics Educators.

Xing, Y. F., Xu, Y. H., Shi, M. H., & Lian, Y. X. (2016). The impact of PM2. 5 on the human respiratory system. Journal of thoracic disease, 8(1), E69.

